# Deep learning-based stratification of Schizophrenia Spectrum Disorder from real-world data reveals distinct profiles of common and rare variant genetic signal

**DOI:** 10.64898/2026.03.30.26349393

**Authors:** Leonardo Cobuccio, Marc Pielies Avellí, Henry Webel, Ricardo Hernandez Medina, Morteza Vaez, Kajsa-Lotta Georgii Hellberg, Yu-Han H. Hsu, Greta Pintacuda, iPSYCH Study Consortium, Anders Rosengren, Thomas Werge, Kasper Lage, Simon Rasmussen

**Affiliations:** Novo Nordisk Foundation Center for Basic Metabolic Research, Faculty of Health and Medical Sciences, University of Copenhagen, Denmark; Institute of Biological Psychiatry, Mental Health Center Sct Hans, Amager and Hvidovre Hospital, Copenhagen University Hospital, Copenhagen, Denmark; Novo Nordisk Foundation Center for Genomic Mechanisms of Disease, Broad Institute of MIT and Harvard, Cambridge, MA 02142, USA; Novo Nordisk Foundation Center for Biosustainability, Technical University of Denmark, 2800 Kongens Lyngby, Denmark; Stanley Center for Psychiatric Research, Broad Institute of MIT and Harvard, Cambridge, MA 02142, USA; Mental Health Center - Sct Hans, Copenhagen University Hospital, Copenhagen, Denmark; The Lundbeck Foundation Initiative for Integrative Psychiatric Research (iPSYCH), Copenhagen and Aarhus, Denmark; Lundbeck Foundation Center for GeoGenetics, GLOBE Institute, University of Copenhagen, Copenhagen, Denmark

## Abstract

Schizophrenia spectrum disorder (SSD) is a clinically and genetically heterogeneous condition, yet few studies have integrated real-world clinical data with both common and rare genetic variation to explore this complexity. In this study, we analyzed real-world data from 22,092 individuals in the Danish iPSYCH cohort (11,046 SSD cases and 11,046 matched population controls) leveraging nationwide registry data on diagnoses, hospitalizations, and parental history. Using a variational autoencoder (VAE), we compressed these features into a latent space and identified ten clinically distinct SSD subgroups that varied in comorbidity, parental diagnoses, hospital burden, and early-life adversity. Polygenic scores (PGSs) for five psychiatric disorders showed subgroup-specific enrichment, highlighting potential links between complex clinical profiles and common variant liability. In a subset with exome data (N=5,969), we assessed rare deleterious variant burden across SCZ-informed gene sets and Protein-Protein Interaction (PPI) networks, observing suggestive network-specific trends. This framework for integrating real world-based stratification with genetic evidence is scalable and transferable across cohorts, offering a path toward biologically informed patient classification.

## INTRODUCTION

Schizophrenia spectrum disorder (SSD) is a genetically complex and highly heritable condition characterized by considerable phenotypic variation and clinical heterogeneity.^1,2^ Although genome-wide association studies (GWAS) have identified many common risk loci,^3–5^ they explain only part of the estimated heritability, which twin studies place at around 80% for Schizophrenia (SCZ) and about 73% for SSD.^6^ Rare variants have also been associated with SSD, with ultra-rare protein-truncating variants in specific genes conferring large increases in SCZ risk (with odds ratios from ∼3 to >20), and may implicate high-impact etiological pathways.^7–9^ Beyond genetics, many environmental and developmental risk factors have been linked to SSD, including prenatal/perinatal exposures,^10^ childhood adversity, cannabis use, migration, urbanicity,^11^ as well as immune-related factors such as infections and autoimmune conditions.^12,13^ This complexity motivates the need to move beyond one-size-fits-all case-control analyses towards exploring clinically meaningful subgroups that may differ in pathophysiology and genetic architecture.^14^ In this scenario, real-world data (RWD), including registries, electronic health records (EHRs), and biobanks, can enable comprehensive clinical and demographic characterization of psychiatric patients and have the potential to advance precision psychiatry.^15–17^

Early biomedical informatics efforts, such as the “Deep Patient” model, showed that unsupervised learning on EHR data could uncover latent patient representations, underscoring the power of high-dimensional clinical data for patient stratification.^18^ More recently, other unsupervised learning approaches have been used to stratify individuals with psychiatric disorders into clinically meaningful subgroups. Multi-Dimensional Scaling (MDS)-based clustering on registry data has revealed distinct diagnostic trajectories in schizophrenia,^19^ and deep learning (DL) has been used to integrate registry and genetic data to identify distinct subgroups across people with schizophrenia and depression.^20^ Additional work has uncovered psychiatric transdiagnostic clusters based on symptom trajectories,^21^ clinical-genetic profiles,^22^ cognitive phenotypes,^23^ and brain connectivity patterns,^24^ illustrating the potential of data-driven stratification across diagnostic boundaries.

Integrating patient stratification with genetic evidence is particularly valuable, as genetically supported drug targets are more than twice as likely to achieve regulatory approval.^25,26^ Large-scale exome sequencing efforts, such as the SCHEMA consortium, have uncovered ultra-rare coding variants in specific genes that confer substantial risk for schizophrenia and converge on synaptic and neuronal processes.^9,27^ When mapped onto brain cell-type-specific protein-protein interaction (PPI) networks derived from human induced neurons, these genetic signals can be interpreted in a functionally grounded, mechanistically rich framework, arguably the closest approximation we have to human neurobiology in disease-relevant contexts.^28–30^ However, it remains unclear whether genetic signals captured in PPI networks can be meaningfully mapped onto clinical subgroups of patients.

Here, we applied unsupervised DL to registry data from 22,092 individuals in the iPSYCH cohort (11,046 SSD cases and 11,046 matched population controls).^31,32^ We identified ten SSD subgroups, capturing differences in comorbidity, hospitalization, developmental profiles, and familial risk. We then tested whether these subgroups differed in their genome-wide polygenic scores (PGSs) for five psychiatric disorders. Additionally, in the subset of individuals with exome sequencing data available (N=5,969; 3,116 cases and 2,853 population controls), we also evaluated differences in the burden of rare, potentially deleterious coding variants across curated schizophrenia gene sets and neuronal PPI modules. We found that specific SSD subgroups exhibited distinct patterns of common genetic liability, with several clusters showing significantly elevated or reduced PGSs for different mental disorders. While rare-variant analyses were underpowered for definitive inference after multiple testing correction, we observed suggestive and directionally consistent patterns in specific clusters. By linking RWD-derived clinical subgroups with both common and, especially, rare genetic signals anchored in strong prior human genetic evidence, we offer a data-driven framework for exploring the genetic underpinnings of clinical heterogeneity. This approach can be extended to other disorders and datasets to uncover biologically grounded dimensions of disease and move toward more refined, genetically informed patient stratification.

## RESULTS

### Study overview

We analyzed data from 22,092 individuals in the iPSYCH cohort,^31,32^ including 11,046 individuals diagnosed with SSD and 11,046 matched population controls (see **Table 1**). We compressed RWD from national registries, including psychiatric diagnoses, hospital contacts, familial risk, and other somatic conditions, into a latent space using a VAE (**Figure 1A**). K-means clustering of these latent representations revealed clinically distinct SSD subgroups. These also differed in their genetic profiles, as reflected by cluster-specific patterns of PGSs and burden of rare predicted deleterious variants across SCZ-related PPI networks, prioritized using Combined Annotation Dependent Depletion (CADD) scores > 15 (see Methods). These findings support a link between clinical heterogeneity and both common and rare genetic variation. To illustrate the rare variant signal underlying these analyses, **Figure 1B** shows the number of CADD variants across the 32 protein coding genes passing FDR correction in the SCHEMA exome sequencing study^9^, together with the total number of alternate alleles observed in SSD cases. More extensive summary statistics aggregated per protein, per network, per dataset and per cluster are available in **Table S1-7** and in **Figure S1**.

**Figure 1.**
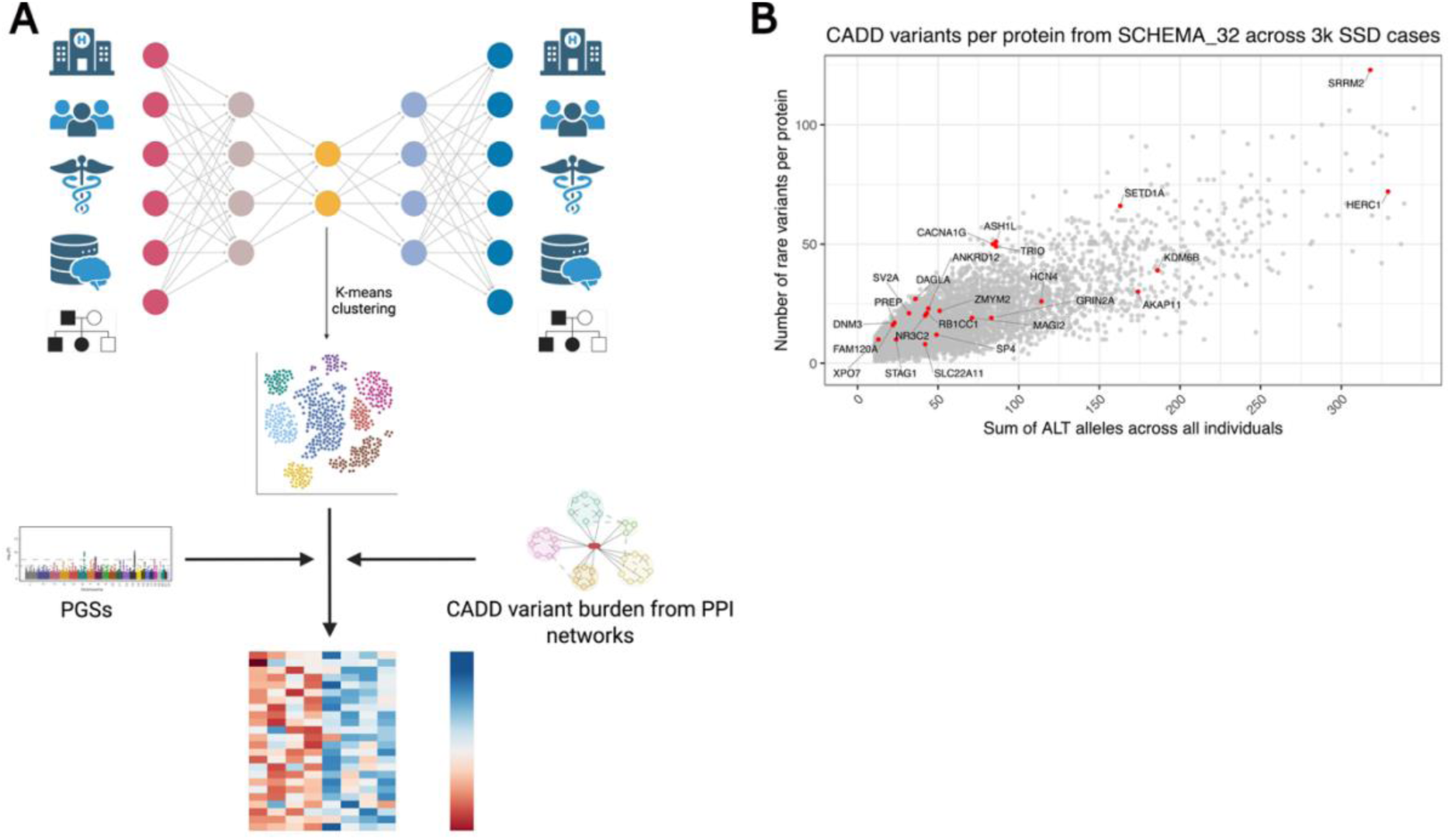
Study overview and distribution of rare variant burden in schizophrenia-associated genes. **A)** Schematic representation of the analytical workflow. We used a VAE on RWD from national health registries, integrating psychiatric and non-psychiatric diagnoses, hospital contacts, and clinical family history on 22,092 individuals with and without a diagnosis of SSD. We then clustered the latent space using k-means to define data-driven subgroups. On top of these clusters, we mapped genetic signal from both common variants (PGSs for psychiatric disorders) and from rare variants (burden of rare variants with CADD score > 15). The rare variant burdens were aggregated across different gene sets, including biologically informed PPI networks. **B)** Scatterplot showing the number of rare CADD > 15 variants per protein (y-axis) and the sum of alternate alleles (x-axis) across all 3,116 individuals diagnosed with SSD and having exome data, focusing on genes from the SCHEMA_32 list. Each dot represents one protein-coding gene, with schizophrenia-implicated genes labeled in red. Proteins such as HERC1, SRRM2, and SETD1A showed higher cumulative variant burden, highlighting genes of potential interest for subtype-specific rare variant analyses.

**Table 1.** Descriptive statistics for all 22,092 individuals, stratified by SSD cases and population controls.

| Variable | Overall, N = 22,092 | Cases, N = 11,046 | Controls, N = 11,046 |
| --- | --- | --- | --- |
| **Has exome data** | 5,969 (27%) | 3,116 (28%) | 2,853 (26%) |
| **iPSYCH1** | 12,035 (54%) | 4,941 (45%) | 7,094 (64%) |
| **iPSYCH2** | 10,023 (45%) | 6,075 (55%) | 3,948 (36%) |
| **ADHD** (attention-deficit hyperactivity disorder) | 1,603 (7.3%) | 1,401 (13%) | 202 (1.8%) |
| **ASD** (autism spectrum disorder) | 1,358 (6.1%) | 1,216 (11%) | 142 (1.3%) |
| **BIP** (bipolar disorder) | 715 (3.2%) | 671 (6.1%) | 44 (0.4%) |
| **MDD** (major depressive disorder) | 4,157 (19%) | 3,760 (34%) | 397 (3.6%) |
| **Age in 2016** |  |  |  |
| Median (IQR) | 27.1 (23.3, 30.9) | 27.1 (23.3, 30.9) | 27.1 (23.3, 30.9) |
| Range | 11.0, 35.7 | 11.0, 35.7 | 11.0, 35.7 |
| **Sex assigned at birth** |  |  |  |
| F | 10,637 (48%) | 5,259 (48%) | 5,378 (49%) |
| M | 11,455 (52%) | 5,787 (52%) | 5,668 (51%) |

**Table 2.** Descriptive statistics for the 5,969 individuals, stratified by SSD cases and population controls, who had exome data available.

| Variable | Overall, N = 5,969 | Cases, N = 3,116 | Controls, N = 2,853 |
| --- | --- | --- | --- |
| **iPSYCH1** | 5,537 (93%) | 2,685 (86%) | 2,852 (100%) |
| **iPSYCH2** | 427 (7.2%) | 427 (14%) | 0 (0%) |
| **ADHD** | 746 (12%) | 644 (21%) | 102 (3.6%) |
| **ASD** | 768 (13%) | 664 (21%) | 104 (3.6%) |
| **BIP** | 304 (5.1%) | 283 (9.1%) | 21 (0.7%) |
| **MDD** | 1,054 (18%) | 997 (32%) | 57 (2.0%) |
| **Age in 2016** |  |  |  |
| Median (IQR) | 26.3 (23.5, 29.6) | 27.1 (24.2, 30.0) | 25.5 (22.7, 28.7) |
| Range | 11.8, 35.7 | 11.8, 35.7 | 13.3, 35.7 |
| **Sex assigned at birth** |  |  |  |
| F | 2,363 (40%) | 1,335 (43%) | 1,028 (36%) |
| M | 3,606 (60%) | 1,781 (57%) | 1,825 (64%) |

### Two distinct clinical profiles emerged from clustering SSD cases and population controls

To assess whether we could differentiate individuals with SSD from unaffected population controls, we applied a VAE to integrate 131 registry-derived features from 22,092 individuals (11,046 SSD cases and 11,046 matched population controls), generating a 40-dimensional latent space after hyperparameter tuning (**Methods**). Feature importance analysis indicated that parental mental disorders and substance use, followed by individual hospital contacts and suicide attempt history, contributed most strongly to the latent structure (**Figure S2A, Table S8**). We then applied k-means clustering on the latent space representations of the individuals in order to stratify them. Clustering validity indices converged on a two-cluster solution as optimal, with the resulting two clusters exhibiting a clear separation (**Figure 2A**): Cluster A was population control-enriched (96.3% of population controls; 10,632/11,046), whereas Cluster B was case-enriched (69.8% of SSD cases; 7,710/11,046), with minimal population control overlap (3.7%). Notably, 30.2% of SSD cases (3,336/11,046) clustered with the population control-enriched group Cluster A, suggesting a subset with milder or less burdened clinical profiles. The case-enriched Cluster B showed a markedly higher clinical burden than the population control-enriched Cluster, with nearly fourfold higher psychiatric diagnosis rates (mean range: 0.012–0.56 vs 0.002–0.17), a 20-fold increase in psychiatric hospital contacts (31.7 vs 1.6 per individual), and substantially higher suicide attempt rates (39% vs 0.01%) (**Figure 2A, Table S9**). Except for Apgar scores, paternal age, and number of previous pregnancies, all differences were statistically significant (Bonferroni corrected p < 0.001 after linear, logistic, and Poisson/negative binomial regressions). Importantly, most features were included as inputs in the VAE model, except smoking during pregnancy, infections and other medical conditions. These were excluded due to limited availability across cohorts, or because information such as birth-related traits may not be consistently available in broader real-world clinical datasets, limiting their applicability for generalizable modeling. The cluster structure hence primarily reflected variation in the modeled clinical features. The clusters also differed on several variables that were not used as input in the VAE model, including higher parental and individual infection burden and more adverse birth-related characteristics for Cluster B. Together, these results show that VAE-derived latent embeddings can capture clinically meaningful structure in real-world registry data, separating a high-burden SSD-enriched subgroup from a lower-burden, population-control enriched group.

**Figure 2.**
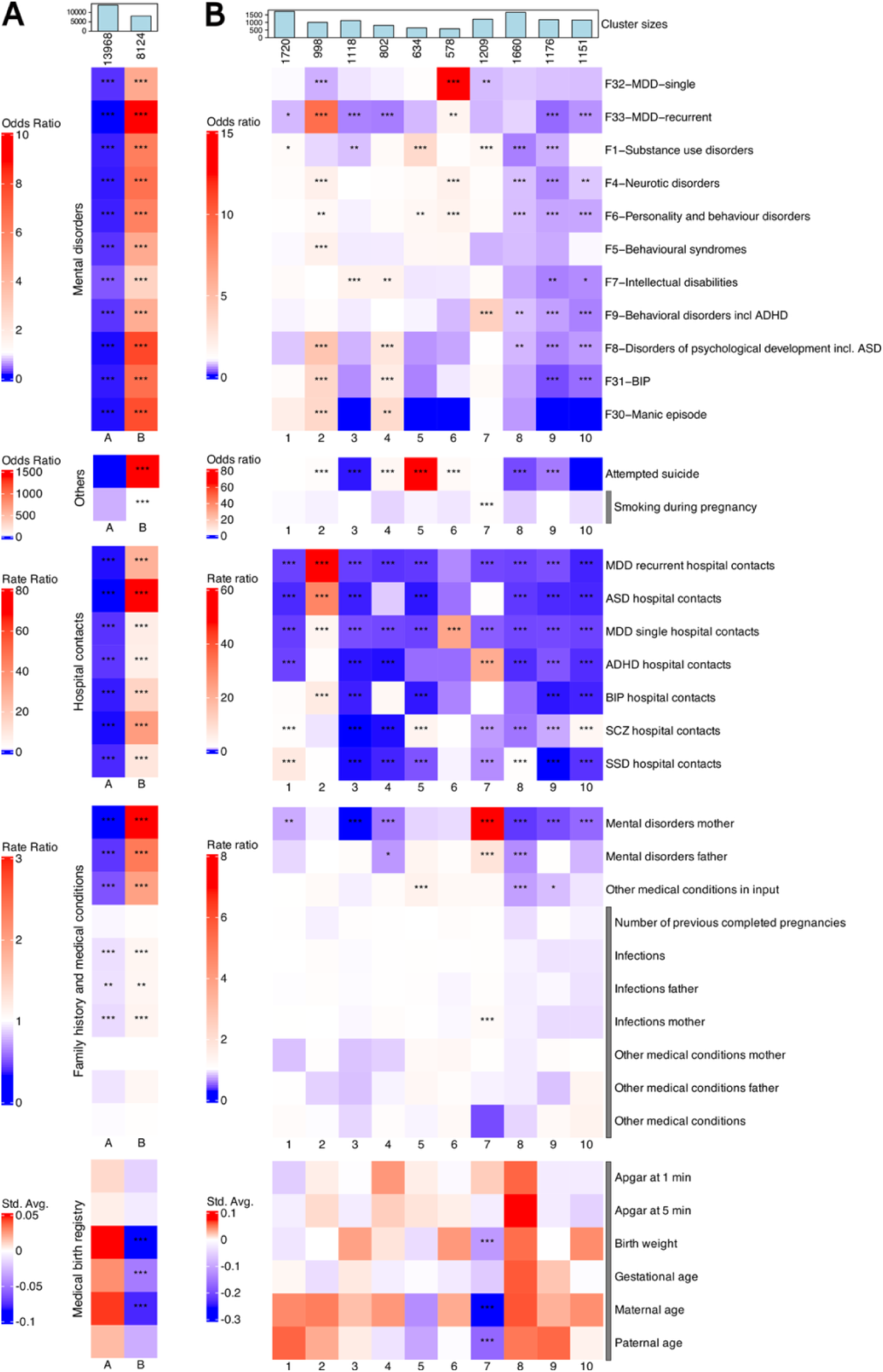
Cluster profiles of individuals with SSD reveal ten distinct clinical subgroups. **A)**Heatmap visualization of the cluster profiles for the 22,092 SSD cases and population controls, divided in clusters A (population controls-enriched) and B (cases-enriched). **B)** Cluster profiles for ten clusters of the 11,046 SSD cases. Mental disorders are grouped by ICD-10 chapter (F1, F4-9) except for the more granular diagnoses of mood disorders: F30-Manic episode, F31-BIP, F32-MDD-single, and F33-MDD-recurrent. Hospital contacts were computed as the sum of all hospital contacts per individual for each main psychiatric diagnosis, i.e. ADHD, ASD, BIP, MDD, SCZ, SSD. The same approach was followed for the remaining count variables, from parental mental disorders to individual other medical conditions. The continuous variables from the medical birth registry were z-score normalized. Color scales reflect: Odds Ratios (ORs) from logistic regressions comparing binary outcomes across each cluster versus all others, Rate Ratios (RRs) from Poisson or negative binomial regressions applied to count data, Standardized averages (Std. Avg.) from linear regressions applied to continuous variables. Unlike beta coefficients, which may obscure subtle trends, standardized averages more clearly highlight cross-cluster patterns. All tests were adjusted for sex assigned at birth, age, iPSYCH1 membership, and the first ten principal components (to account for potential residual population stratification). The significance levels are defined as: p-value < 0.05: *, p-value < 0.01: **, p-value < 0.001: *** (all after Bonferroni correction). The variables with a grey bar next to their labels were not used as input features for the VAE model.

### SSD cases stratify into ten subgroups, seven of which show severe comorbidities

We then proceeded to identify more fine-grained structure in the SSD cases. To this end, we applied k-means clustering to the latent representations from a VAE retrained on the 11,046 SSD cases. This latent space was primarily driven by parental mental disorders, particularly substance abuse, in addition to individual hospital contacts (**Figure S2B, Table S10**), and revealed ten clinically distinct groups (**Figure 2B** and **Table S11** for all test p-values and effect sizes). Clusters 1 through 7 captured more severe and comorbid subgroups, though with differing clinical signatures. Cluster 1 was marked by elevated rates of substance use diagnoses, along with high SCZ-related hospital contacts and the highest hospitalization rate for SSD (Rate Ratio 6.42; average count = 114; p < 0.001, Poisson regression), but lower number of recurrent MDD diagnoses. Cluster 2 was the most comorbid, spanning multiple mood, anxiety, and developmental disorders, in addition to having the highest rates of recurrent MDD (OR 9.335, avg 0.495, p-value < 0.001, logistic regression), and the greatest hospital contact burden for recurrent MDD, ASD, and BIP. Clusters 4 and 6 ranked next in overall comorbidity after Cluster 2, with Cluster 4 showing enrichment for intellectual disability, developmental disorders, bipolar disorder, and manic episodes, and Cluster 6 marked by elevated rates of major depressive disorder and neurotic, personality, and behavioral disorders.

Other clusters showed more specific profiles, including high rates of intellectual disability but low suicidal behavior (Cluster 3), and elevated substance use and suicidal behavior alongside more non-psychiatric medical conditions (Cluster 5). Cluster 7 stood out for having higher substance use, behavioral disorders including ADHD, and ADHD hospital contacts, along with increased exposure to early-life adversities such as maternal infections and low birth weight, in addition to having younger parents. Together, these findings illustrate how unsupervised clustering of real-world registry data can reveal clinically meaningful subgroups within SSD, underscoring its clinical heterogeneity, and sets the stage for exploring their distinct genetic underpinnings.

### Three SSD subgroups showed reduced clinical severity and may reflect stable or focused psychotic profiles

Clusters 8-10 reflected less severe clinical profiles of SSD cases when compared to clusters 1- 7. These groups had markedly lower psychiatric comorbidity, hospital contacts, maternal mental illness, and suicidality. More specifically, Cluster 8 showed reduced hospital contacts across most disorders, including SCZ, but maintained higher SSD-related hospital use. This discrepancy suggests that psychotic presentations in Cluster 8 are less frequently classified as schizophrenia (F20) and more often fall within other psychotic-spectrum diagnoses (F21–F29). Cluster 9 had the lowest rates of psychiatric diagnoses and hospital contacts across all conditions. These individuals may represent stable or less severely affected cases, or potentially people whose diagnosis may have changed or improved over time. Cluster 10 also had lower rates of co-occurring psychiatric disorders, but average rates for substance use disorders and behavioral syndromes. However, it had higher numbers of hospital contacts for SCZ, suggesting a form of SSD that was more narrowly focused on psychotic symptoms without other major mental health issues.

### Strong PGS enrichments mirror clinical severity and neurodevelopmental factors

To explore whether clinical heterogeneity within SSD reflected underlying differences in common genetic liability, we examined how the clinically defined clusters differed in terms of polygenic scores (PGSs) for five psychiatric traits: attention-deficit hyperactivity disorder (ADHD), autism spectrum disorder (ASD), bipolar disorder (BIP), major depressive disorder (MDD), and schizophrenia (SCZ). As expected, the case-enriched Cluster B had higher PGSs across all five psychiatric traits compared to the population control-enriched Cluster A (**Figure 3A left, Table S12)**. Conversely, the PGS profiles across the ten SSD clusters revealed heterogeneous patterns.

**Figure 3.**
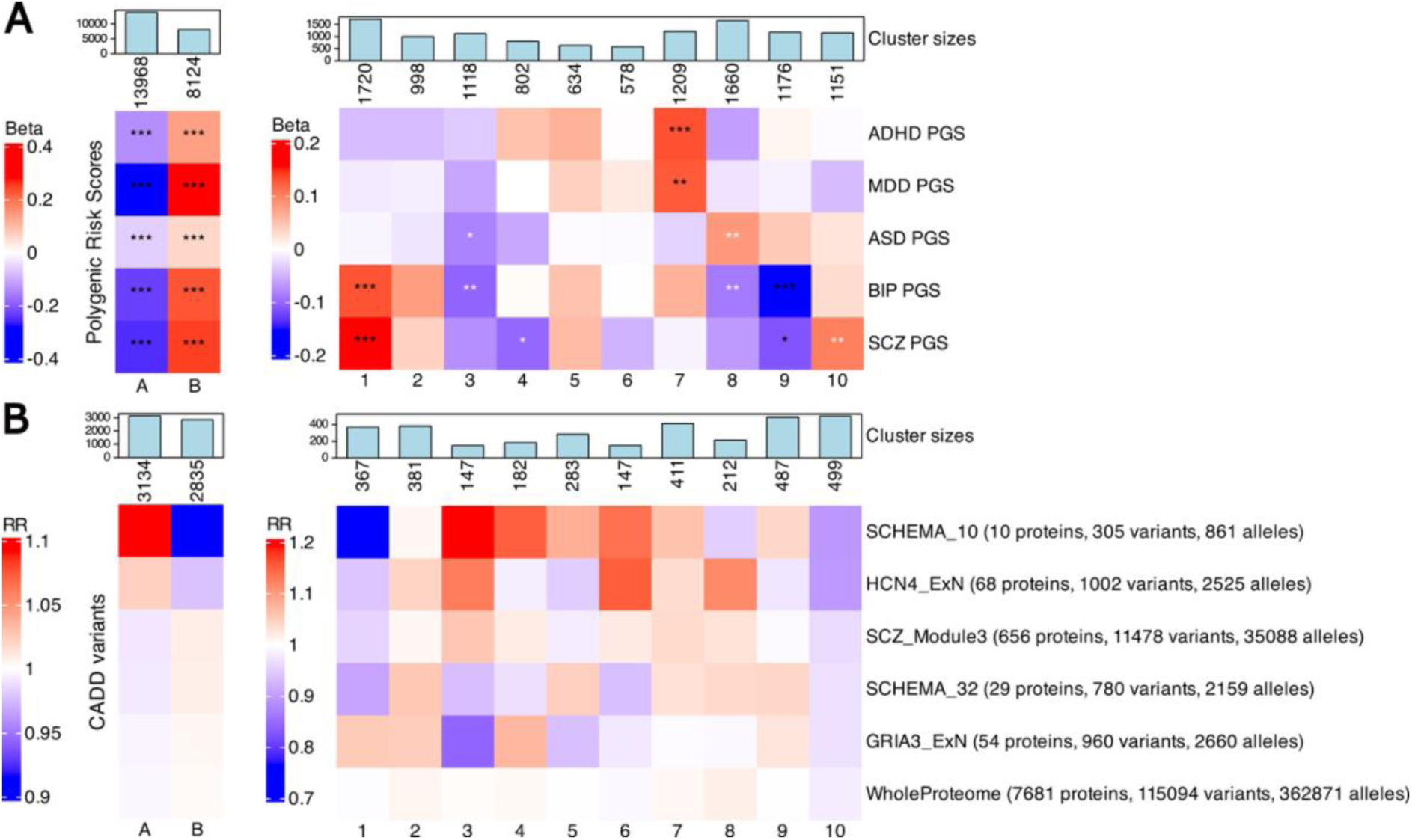
Distinct genetic profiles across clinically derived SSD subgroups. **A)** Heatmaps showing the results of the association analyses for genome wide PGSs of five psychiatric disorders, ADHD, MDD, ASD, BIP, and SCZ, across the initial population control vs. SSD case clustering (left) and the ten SSD clinical subgroups (right). Beta coefficients are derived from linear models comparing each cluster to all others, adjusted for sex assigned at birth, age, first ten principal components, and iPSYCH1 membership. Significant enrichments are indicated by black asterisks (after Bonferroni correction) and white asterisks (after FDR correction): p-value < 0.05: *, p-value < 0.01: **, p-value < 0.001: ***. **B)** Heatmaps showing rate ratios (RRs) of rare CADD >15 variant burden across curated schizophrenia gene sets and protein–protein interaction (PPI) networks. Left panel shows comparison between SSD cases and population controls; right panel displays burden across the ten SSD subgroups.

Cluster 1, defined by high SSD and SCZ related hospital contacts and high substance use, showed the highest levels of PGSs for both SCZ and BIP (with betas of 0.158 and 0.135, and standardized averages of 0.135 and 0.131, respectively, both with Bonferroni p-value < 0.001) (**Figure 3A left, black asterisks**, **Table S13**). This supported a potential link between SSD symptoms severity and common variant burden for SCZ and BIP. Cluster 7, which had higher behavioral disorders diagnoses, ADHD hospital contacts, and early-life adversity, showed significantly elevated PGSs for ADHD and MDD, pointing to shared common polygenic liability across neurodevelopmental traits and behavioral dysregulation. Individuals in Cluster 9, characterized by the lowest comorbidity and hospital contacts across all disorders, showed significantly lower PGSs for BIP and SCZ, reaching values of standardized averages of -0.132 for BIP (Beta -0.154, p-value < 0.001) and of -0.092 for SCZ (Beta -0.114 p-value < 0.05). This could potentially indicate a less genetically loaded presentation of SSD.

### Modest PGS enrichments hint at alternative or non-genetic risk in specific SSD groups

Other clusters showed more modest patterns, reaching significance only after FDR correction. Cluster 3, defined by developmental delay and low overall hospital burden, was the only cluster with consistently lower PGSs across traits, reaching significance for ASD and BIP (**Figure 3A right, white asterisks, Table S13**). This suggested a potentially greater contribution of non-polygenic factors, such as environmental influences or rare variants. Cluster 8, which had low overall comorbidity but maintained SSD-related hospital contacts, showed higher ASD PGS and lower BIP PGS, potentially reflecting a distinct liability profile in which core neurodevelopmental genetic risk manifests without broader mood or behavioral comorbidity. Cluster 10 was the second cluster, after Cluster 1, to have significantly higher PGS for SCZ (after FDR correction, Beta 0.102, std avg 0.105, q-value 0.0089). Given its high SCZ-related hospital contact burden and limited comorbidity, this pattern is consistent with an SSD presentation predominantly centered on psychotic features and enriched for schizophrenia-specific polygenic liability. Overall, these findings show that PGSs for psychiatric disorders were unevenly enriched across SSD subgroups, reflecting specific clinical dimensions such as psychosis severity, neurodevelopmental features, and comorbidity, highlighting the value of stratified analyses to capture SSD heterogeneity.

### Case-enriched cluster shows reduced rare variant burden in SCHEMA_10, unlike broader SCZ gene lists

Next, we assessed whether clinical subgroups differed in their number of rare CADD variants aggregated across SCZ related gene lists and PPI networks (see Methods). Although we could not detect any statistically significant signal, the distribution of rare CADD variant burden across gene sets revealed suggestive trends. The first pattern that stood out was related to the SCHEMA_10 gene set, comprising the ten genes most strongly associated with schizophrenia via ultra-rare coding variants in the SCHEMA study.^9^ In our case vs population control-enriched analysis, this gene set showed a trend in the opposite direction: individuals in the case-enriched Cluster B had a slightly lower burden of CADD variants in these genes compared to the population control-enriched Cluster A (RR 0.915, avg 0.271 vs 0.294) (**Figure 3B left, Table S14**). A similar pattern was observed for the HCN4_ExN protein network.

Notably, all other protein lists and particularly SCHEMA_32, a broader gene set that contains all SCHEMA_10 genes, showed a reversed direction of effect, with slightly higher CADD variants burden in the case-enriched Cluster B. This divergence may reflect statistical noise due to the small number of genes in SCHEMA_10 and HCN4_ExN (10 and 68, respectively).

### CADD variant burden from SCZ-informed PPI networks show suggestive patterns across SSD clusters

This further motivated a deeper examination of the CADD burden in the within-case stratification. Here we observed that the burden of CADD variants in the SCHEMA_10 was lowest in Cluster 1, which had the most severe hospital contact profile for SCZ and SSD (**Figure 3B right, Table S15**). Conversely, the highest burdens for SCHEMA_10 were found in clusters 3 and 4, characterized by developmental or mixed comorbidity profiles, and not by SSD severity per se. The pattern of CADD burden across the 10 SSD clusters of the protein networks HCN4_ExN and SCZ_Module3, PPI modules identified in stem-cell derived neurons from SCZ associated genes, most closely resembled that of SCHEMA_10 (Pearson correlation coefficient of 0.52 and 0.75, respectively), with opposite direction effects for Cluster 5 and 9 (positive in SCHEMA_10, negative in the others). GRIA3_ExN, a PPI network centered on GRIA3, was the gene list that differed the most from the SCHEMA_10, HCN4_ExN, and SCZ_Module3, with correlations coefficients of -0.42, -0.36, -0.39, respectively. It was also the only protein network with higher CADD variants burden in Cluster 1, in addition to Cluster 2 and 4, while it had a lower burden in Cluster 3 and 5 compared to the rest of the clusters. Interestingly, Cluster 10, one of the least comorbid clusters, but characterized by higher hospital contacts related to SCZ diagnosis, was the only one having a lower CADD burden across all 6 gene lists. Finally, as expected, the WholeProteome background set, a list of proteins expressed in stem cell derived neurons, detected by mass spectrometry, showed the least amount of variation across both the first clustering (**Figure 3B left**), and across the 10 SSD clusters (**Figure 3B right**), with average number of CADD variants per cluster ranging from 115 to 117, compared to the overall average of 116. Together, these findings underscore the importance of integrating clinical heterogeneity when interpreting genetic signals, even from high-confidence gene sets. While preliminary and not statistically significant, these patterns suggest that rare variant load may not be uniformly elevated across all SSD presentations and may instead concentrate in more narrowly defined subgroups.

## DISCUSSION

Connecting molecular risk signals with the clinical heterogeneity in schizophrenia spectrum disorders (SSDs) remains a central translational challenge in psychiatric genetics. Although GWAS and exome sequencing have identified many risk loci and rare variants for schizophrenia,^5,9,33^ these findings have yet to be effectively integrated with real-world clinical data to inform patient stratification. Here, we stratified 22,092 individuals with and without SSD from the iPSYCH cohort using unsupervised clustering of clinical registry data integrated through a DL model. We then mapped genome-wide polygenic scores for psychiatric disorders and rare coding variant burden from biologically informed gene sets and PPI networks across the resulting subgroups. This extends our previous work^20^ to more individuals with SSD and to focus on mapping of rare genetic signal.

We identified ten clinically distinct SSD subgroups with varying comorbidity, early-life adversity, hospitalization profiles, and family history. Clusters 2 and 4 were both highly comorbid, but only Cluster 2 showed elevated hospital contacts, possibly reflecting differences in access to care or disease course. Clusters 8–10 showed the lowest overall burden, while Cluster 7 stood out for its neurodevelopmental profile: lower parental age, low birth weight, high ADHD comorbidity, and familial psychiatric history, suggesting a distinct early-life risk profile. This stratification aligns with prior evidence that schizophrenia is not a homogeneous disorder but includes subgroups defined by differences in cognitive trajectories and premorbid functioning, including neurodevelopmental and affective profiles.^34,35^

These clinical differences were mirrored by distinct patterns of common genetic risk. Cluster 1, marked by substance use and high hospitalizations, had the highest PGSs for SCZ and BIP, possibly suggesting that their severity may, in part, be associated with or potentially driven by an elevated burden of common risk variants. Cluster 7, with neurodevelopmental traits, had elevated PGSs for ADHD and MDD. Conversely, Cluster 3, defined by developmental delay and low clinical burden, had uniformly low PGSs, pointing to alternative etiologies, including rare or environmental factors. These results are consistent with earlier research suggesting that PGSs can differentiate SSD subgroups by cognitive course, affective comorbidity, and age at onset,^36,37^ and that common genetic variation maps onto clinically meaningful dimensions such as cognition, externalizing behavior, and mood instability.^38^ Although PGSs have limited predictive power for psychiatric disorders,^39^ they may still capture clinically meaningful variation across symptom dimensions, potentially helping in understanding the etiological differentiation of clinical subtypes.

A key contribution of our study is the mapping of rare coding variant burden (filtered for CADD score > 15 to prioritize variants with likely functional impact) from biologically informed SSD-relevant gene sets and PPI networks. These networks represent curated modules derived from orthogonal molecular and *in silico* experiments spanning multiple omics layers and offer a biologically grounded scaffold for grouping rare variants with potentially shared functional consequences towards schizophrenia.^30^ While no comparisons reached statistical significance after multiple testing correction, suggestive patterns emerged. SCHEMA_10 and HCN4_ExN showed lower rare variant burden in the case enriched Cluster B than in the population control-enriched Cluster A, unlike broader gene sets such as SCHEMA_32 **(Figure 3B)**. While this may reflect limited power due to small gene sets and sample size, it might also be consistent with Simpson’s paradox, where aggregate effects mask or even reverse trends present within subgroups. Indeed, within the SSD cases, the SCHEMA_10 and HCN4_ExN gene sets showed lower burden in Clusters 1 and 10 compared to other gene sets. These subgroup-specific depletions may have disproportionately influenced the first clustering (Cluster A vs B), creating an apparent depletion in Cluster B that does not reflect the broader distribution within the cases. Cluster 3, notable for its low PGSs and developmental profile, had a higher burden for the SCHEMA_10 and HCN4_ExN gene sets. This cluster may represent a SSD subgroup more influenced by deleterious variants in these gene sets, consistent with evidence that schizophrenia and neurodevelopmental disorders share pathogenic coding variants, suggesting that certain schizophrenia subtypes might lie on a continuum of neurodevelopmental impairment.^40^ This cluster, however, had lower rare variant burden for SCHEMA_32. Such seemingly counter-intuitive findings reinforce the need to account for clinical heterogeneity when interpreting genetic burden, even in gene sets with strong prior associations. In contrast, Clusters 1 and 10 had low rare variant burden across almost all gene sets, but high SCZ PGSs, aligning with the notion that rare and common variant contributions may inversely correlate. GRIA3_ExN burden was elevated in highly comorbid Clusters 2 and 4, suggesting that different biological modules may track distinct aspects of clinical complexity. Together, these results suggest that rare variant burden within SCZ-informed PPI networks is unevenly distributed across SSD subgroups may help link distinct genetic signals to clinical phenotypes. Prior systems genomics, multi-omics, and stem-cell studies have shown that schizophrenia risk genes converge on synaptic and neurodevelopmental pathways, supporting the biological relevance of these network-based analyses^7,41,42^

While exploratory, our results underscore the potential value of integrating rare genetic data with real world-based patient stratification to help improve our understanding of psychiatric disease mechanisms. Larger samples and improved annotations will be key to validating these early signals. More broadly, our approach supports the view that distinct routes to schizophrenia may exist, some shaped by polygenic risk, others by rare variants or early adversity.^16^ Recognizing compensatory or divergent genetic architectures across subgroups may help resolve inconsistencies across studies or patient cohorts and could ultimately inform more precise interventions.

Our findings are situated at the intersection of two major research currents in psychiatry: (1) the movement toward biologically informed nosology, and (2) the rise of multi-modal and network-based approaches to elucidate disease mechanisms. The need to move beyond DSM-based diagnostic categories has been repeatedly emphasized,^43,44^ and newer efforts have sought to redefine mental disorders using dimensional, data-driven approaches.^45–47^ Our latent clustering of registry data contributes to this goal by revealing clinically coherent subgroups that cut across traditional diagnostic boundaries, with early indications of validity supported by their alignment with genetic risk profiles. Finally, by using real-world registry data, researchers can leverage information that is increasingly available in many healthcare systems and biobanks. Combined with genetic approaches such as PGSs and network-informed rare variant mapping, our approach could be applied to other complex traits with known rare variant contribution, including mood disorders, autism, or non-psychiatric diseases marked by phenotypic and genetic heterogeneity, such as diabetes, obesity, and cardiovascular disease.^48–51^ In particular, many nonpsychiatric disorders are studied in far larger cohorts and therefore offer substantially greater power for rare variant analyses.

## LIMITATIONS

This study has several limitations. First, although national health registries contain rich longitudinal data, our chance to model this complexity was constrained by the low number of variables available in our setting, i.e. 131 input variables. Second, while unsupervised DL enables discovery of latent clinical structure, it poses challenges for interpretability and replication. The subgroups we identified were data-driven and post hoc, requiring external validation before firm conclusions can be drawn. A further limitation concerns the size and composition of the genetic dataset. While the full clinical cohort was relatively large, the exome sequencing data was available only for a smaller subset, reducing the power of our rare variant analyses. Another limitation concerns temporal changes in diagnostic practices. During the study period (1980-2008), recognition of conditions such as autism and ADHD increased substantially, and thus individuals born earlier in the cohort were less likely to receive these diagnoses, which in turn may influence diagnosis-based features and the clinical patterns captured by the model. In addition, some gene sets used, such as SCHEMA_10, SCHEMA_32, and HCN4_ExN, were small and prone to high variance. Finally, the SCHEMA-derived gene sets and PPI networks we used in this study were derived using datasets that included individuals from our own iPSYCH cohort, potentially introducing circularity and an inflation of associations. However, given the exploratory nature of our analyses and the lack of statistically significant rare variant findings, our results do not constitute new genetic associations but rather illustrate the potential of combining clinical subtyping with biologically informed gene sets as a framework for future studies.

## MATERIALS AND METHODS

### Dataset

We used the iPSYCH2015 dataset from the Danish iPSYCH consortium,^31,32^ covering all individuals born in Denmark between 1981-2008 (N = 1,657,499). From this population, 88,831 psychiatric cases and 48,420 population-based controls were genotyped, yielding a case-cohort sample of 137,251 individuals. The cohort was assembled in two stages: iPSYCH1 (known as iPSYCH2012, with 57,377 cases and 30,000 population controls), and iPSYCH2 (known as iPSYCH2015i, with 36,741 cases, 19,982 population controls). DNA was collected at birth from dried blood spots stored in the Danish Neonatal Screening Biobank.^52^ Genotyping used the Infinium PsychChip v1.0 for iPSYCH1, and the Global Screening Array v2 for iPSYCH2. For both arrays, stringent quality control and imputation pipelines were applied as described in Schork et al. 2019.^53^ Among the 22,092 SSD cases and population controls included in this study, 5,969 individuals (belonging to the iPSYCH1 wave) had exome data that was generated in Singh et al. 2022^9^ and that was processed following Satterstrom et al. 2019.^54^ Only unrelated participants of European ancestry were retained for the analysis. Diagnoses were retrieved through the Danish Psychiatric Central Research Register^55^ and the Danish National Patient Register.^56^ These registries capture all specialist-led psychiatric assessments conducted in both inpatient and outpatient settings and did not include data for primary care contacts. Diagnoses were recorded under the International Classification of Diseases, Eight Revision or Tenth Revision (ICD-8, ICD-10), and included attention-deficit/hyperactivity disorder (ADHD: F90.0 ICD-10 code), autism spectrum disorders (ASD: F84.0, F84.1, F84.5, F84.8, F84.9), bipolar disorders (BIP: F30-31), major depressive disorder (MDD: F32, F33), schizophrenia spectrum disorder (SSD: F20-F29). For this study, we included all SSD individuals as cases, totaling 11,046. We then extracted an age-matched sample of 11,046 individuals from the random population, not having been diagnosed with SSD, to use as population control. The identities of all participants were pseudonymized in compliance with data protection regulations. The study protocol was approved by the Danish Data Protection Agency, and, under Danish law, individual informed consent was not required. For the VAE analysis, we compiled a dataset of 131 variables derived from registry data, which captured both the clinical and familial aspects of patient health. These variables included age-at-diagnosis information for all psychiatric diagnoses recorded in the registry available (ICD-10 codes from F00 to F99), as well as some non-psychiatric diagnoses that were available. Additionally, we included severity measures such as attempted suicide and number of hospital contacts related to psychiatric diagnoses. Finally, we included binarized parental psychiatric diagnoses and some non-psychiatric related diagnoses as a family history indicator. Other variables were not included in the input data. For example, although birth registry information was available, this was excluded from the input data as we deemed it to be not actionable within the bigger scope of this proof-of-concept study. Moreover, we didn’t include infections and autoimmune diagnoses as these were generally available only for individuals in the iPSYCH1 cohort. Nevertheless, we incorporated these external variables into the cluster profile analysis to provide additional insight into the identified subgroups. The complete list of all variables is available in **Table S16**.

### Preprocessing of input data to VAE model

Continuous variables, including age at diagnosis and the number of hospital contacts, were standardized using z-score normalization after training and test split, resulting in a mean of zero and unit variance for each feature. Missing values in these continuous measures were replaced with the feature’s mean value (zero after normalization). Categorical variables, such as family history and suicide attempts, were one-hot encoded and subsequently flattened for model input, with absent values represented by an all-zero vector. To ensure data robustness, only variables observed in at least 1% of cases were retained for analysis. After this filtering step, 131 variables were included as input data to the VAE model. See **Table S16** for an overview of all variables.

### VAE implementation and model

We integrated the multimodal data using a β-Variational Autoencoder (VAE) model, adapted from the MOVE framework.^57^ The model consisted of fully connected encoder and decoder networks with LeakyReLU activations,^58^ batch normalization, and dropout rate of 0.2, mapping inputs to a Gaussian latent space via the reparameterization trick. Reconstruction losses were computed separately for continuous and categorical variables using cross-entropy and mean squared error, respectively. The overall loss combined these reconstruction terms, which were normalized by dataset size and batch size, with a Kullback-Leibler divergence (KLD) term that regularized the latent space towards a standard normal distribution, weighted by a tunable parameter β. We defined the loss as:

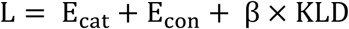

Where E_cat_ and E_con_ are vectors of the reconstruction error for the categorical and continuous datasets, β is the weight applied to the KL-divergence KLD. Following Kingma et al, 2013,^59^ this was defined as:

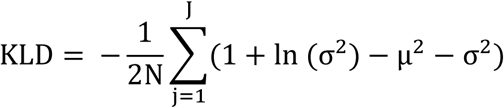

where N is the batch size, and the summation is performed over the J latent dimensions for each sample. Missing continuous data were encoded as feature-wise mean values (zero after z-score normalization) and excluded from back-propagation, while missing categorical data were represented as zero vectors and handled via an ignore index in the loss computation to not be backpropagated. The VAE was optimized using the Adam optimizer,^60^ and the latent representation for each individual was derived by extracting the mean vector from the encoder after training. During training, we implemented a dynamic schedule to improve convergence. In particular, the batch size was dilated at predefined epochs 50, 100, and 150 by a factor of 1.5 to gradually incorporate more samples per mini-batch. We set the final number of training epochs to 300 to allow the model to train for a considerable amount of time. Additionally, we ran the training using a KLD-warmup strategy, in which the KLD weight β was set to 0 for the first epochs, and then incrementally raised from epoch 80 through epoch 150 until it reached its final chosen value of 0.001 (see Hyperparameter tuning). This approach allowed for a gradual regularization of the latent space while the model adjusted to increasing batch sizes over the course of training.

### Hyperparameter tuning

We randomly selected 11,000 individuals from the full dataset (N = 22,092) and split them into 80% training and 20% test sets using stratified sampling. Continuous variables were standardized after the split to prevent data leakage. We performed a grid search to optimize the variational autoencoder (VAE), varying the number of hidden layers (1, 2, or 3), the batch size (32, 64, or 128), the number of latent dimensions (2, 10, 20, 30, or 40), and the β coefficient of the Kullback–Leibler divergence (KLD) term (0 to 10 across 14 values, see **Figure S3, Table S17**). KLD warm-up schedules were applied, gradually increasing β during initial epochs. We evaluated models using reconstruction error (cosine similarity for continuous variables, top-probability match for categorical ones) and test log-likelihood, excluding missing data. Based on these results, we selected 0.001 as the best β value, representing the point just before train and test reconstruction performances started to degrade (**Figure S4**). Among all models trained with β = 0.001, we ranked the top 10 by aggregate performance across reconstruction (train/test) and test log-likelihood (**Figure S5B, Table S18**). To assess stability, each model was retrained 10 times and compared via changes in cosine similarity across latent projections. The model with the smallest average difference was deemed most stable (**Figure S6, Table S19**). The final model used one hidden layer of 600 neurons, a batch size of 64, 40 latent dimensions, and β=0.001, and was used to generate latent representations for both the full dataset (N = 22,092) and the SSD cases-only subset (N = 11,046).

### Feature importance on latent space

We quantified how individual input features influenced the latent space learned by the VAE by employing a perturbation-based feature importance method, as implemented in MOVE.^57^ For each categorical and continuous feature, we generated perturbed datasets in which the target feature was set to missing, represented by an all-zero vector for categorical features and a value of zero for continuous ones, while keeping all other inputs unchanged. Each perturbed dataset was passed through the trained model to obtain new latent embeddings. We then measured the per-sample difference between the original and perturbed embeddings by summing the absolute changes across latent dimensions. The absolute differences in the latent representations were then aggregated across samples to yield an overall impact score per feature (**Figure S2**, **Table S8 and S10**).

### Clustering

To assess whether the VAE-derived latent space contained meaningful clustering structure, we computed the Hopkins statistic, a measure for assessing the spatial randomness of multivariate data, using the `hopkins` function from the clusterend R package v1.7.^61^ The result (Hopkins of 0.0515 for the SSD cases and population control dataset, and of 0.1235 for the SSD cases only) indicated strong deviation from randomness and the presence of underlying cluster structure. For the combined dataset of SSD cases and population controls, we used the R package NbClust v3.0.1 with 26 clustering indices to estimate the optimal number of clusters.^62^ As expected, the algorithm supported number of clusters k = 2, reflecting diagnostic separation between cases and population controls (**Figure S7A, Table S20**). Clustering the cases-only dataset posed a greater challenge, because no clear ground truth was available and clustering algorithms may identify spurious structure in high dimensional data. We thus we adopted a more robust two-step procedure combining clustering indices from NbClust and Monte Carlo Reference-based Consensus Clustering (M3C) to assess their statistical significance by comparing them to a null reference distribution generated.^63^ NbClust supported k = 2, 6, and 10 (**Figure S7B, Table S20**), but only k = 10 yielded a statistically significant M3C p-value (p < 0.05; **Figure S7C-D, Table S21**). Thus, we selected k = 10 as the final number of clusters for downstream analyses.

### Consensus clustering and clusters stability

To ensure robust clusters, we ran k-means clustering 100 times on the VAE-derived latent space using the `kmeans` function from base R with identical parameters. We then built a consensus matrix capturing how often each pair of individuals was clustered together, reducing sensitivity to random initialization. Final clusters were derived by reapplying k-means to this consensus matrix. To assess clustering stability, we computed Adjusted Rand Index (ARI) values across all pairs of runs using the `cl_agreement` function from the clue package v0.3.65.^64,65^ The Rand index quantifies the agreement between two clusterings by measuring the proportion of sample pairs consistently grouped together (or apart) across runs. The ARI, ranging from 0 (no agreement) to 1 (perfect agreement), had a median of 0.46 for the full SSD cases and population control dataset and 0.56 for the SSD-only latent space (**Figure S8, Table S22**).

### Data preprocessing for cluster profile analyses

To perform the cluster profile analysis (**Figure 2**), we aggregated binary, count-based and continuous variables from registry data (see **Methods: Dataset**). For each individual, we grouped psychiatric diagnoses by ICD-10 chapters F1 and F4–F9, with each chapter collapsed into a binary variable (1 = any diagnosis present, 0 = absent). Due to their prevalence and clinical relevance, we retained the ICD-10 diagnoses F30 (manic episode), F31 (bipolar disorder), F32 (single depressive episode), and F33 (recurrent depressive disorder) as separate binary variables. Attempted suicide and maternal smoking during pregnancy (the latter not used as input in the VAE model) were encoded as binary variables. We aggregated hospital contact variables, which included number of contacts and total days admitted, for each person and for each key psychiatric diagnostic category: ADHD, ASD, BIP, SCZ, SSD, and MDD (both single and recurrent episodes).

Parental psychiatric diagnoses were summarized by counting ICD-10 F-chapter diagnoses per parent. Thirteen somatic conditions (e.g., asthma, epilepsy, congenital anomalies) present in ≥1% of cases were summed into a single “Other medical conditions in input” variable to provide a cumulative indicator of somatic comorbidity beyond psychiatric diagnoses. For infections, we applied the same approach by summing the number of infection-related diagnoses for each individual and their parents. These variables were excluded from the VAE model input, as they were predominantly available only for individuals in the iPSYCH1 cohort. We aggregated a set of 30 non-psychiatric autoimmune and inflammatory conditions (**Table S16**), including, for example, type 1 diabetes, ulcerative colitis, rheumatoid arthritis, multiple sclerosis, and celiac disease, and counted how many of these diagnoses were present for each individual and for each of their parent. Again, these were not used as input to the VAE model. Finally, continuous variables from the birth registry (not used in VAE input) were z-score normalized before cluster analysis.

### PGS computation

To generate the PGSs, we obtained the latest external GWAS summary statistics for ASD,^66^ ADHD,^67^ SCZ,^5^ MDD,^68^ and BIP^69^ provided by the Psychiatric Genomics Consortium (PGC)^70^ in which the iPSYCH2015 individuals were excluded from the discovery sample of GWASs. After intersecting the SNPs in the target sample (i.e., iPSYCH2015) with the SNPs in HapMap3 as the reference, 1,041,920 SNPs were retained for PGS computation. The p-value threshold was set at 0.9 to determine SNP inclusion. To rescale the SNP effect sizes in each summary statistic, we used SBayesR^71^, which applies Bayesian multiple regression. The analysis was performed with default parameters for the number of mixture components (π) and the scaling factor (γ), specified as *--pi* 0.95,0.02,0.02,0.02,0.02,0.01 and *--gamma* 0,0.01,0.1,1, respectively. To compute the PGSs, we applied the *--score* module in PLINK v2.00a2.3LM to the rescaled SNPs’ effect sizes derived from SBayesR output files. The scores were subsequently normalized by subtracting the mean score of the control population (i.e., the iPSYCH2015 random cohort) and dividing by the standard deviation of scores within the control group.

### Exome data processing

All variant calls (VCF files) were originally aligned to the hg19 (GRCh37) reference genome. For consistency with more recent assemblies, these were lifted over to the GRCh38 reference using bcftools (v1.20)^72^ with the `+liftover` plugin^73^ to each chromosome-level VCF while supplying the hg19ToHg38.over.chain.gz chain file downloaded from the UCSC Genome Browser.^74^ The VCF files were then sorted with `bcftools sort` and normalized through `bcftools norm` (with the `-m -any` option) to split multiallelic records and left-align INDELs. This pipeline step also removed any duplicate entries, yielding one final VCF file per chromosome in compressed and indexed format. All lifted and normalized files were subsequently annotated with Ensembl Variant Effect Predictor (VEP; version 112),^75^ which provided information on Combined Annotation–Dependent Depletion (CADD) scores to prioritize potentially deleterious variants.^76^ CADD scores integrate multiple annotation metrics into a single value, with higher scores indicating an increased likelihood that a variant is deleterious and adversely affects protein function.^76^ We filtered the exome data to retain only rare variants (< 1% AF) with CADD score > 15, a widely used cutoff for identifying variants likely to impair protein function and which coincides with the median score for canonical splice site and missense variants.^77^ For each individual, we counted the number of alternate alleles located in the coding regions of each protein-coding gene. We then summed these allele counts across genes belonging to predefined gene and protein sets, defined the following section. We used the resulting value, the sum of CADD alternate alleles per gene set per individual, to perform enrichment testing for each cluster against the rest (see **Method: Statistical tests**).

### Derivation of gene sets

To investigate rare variant burden within biologically meaningful contexts, we analyzed curated gene sets derived from large-scale genetic association studies and neuronal protein–protein interaction (PPI) networks linked to schizophrenia. From the SCHEMA exome sequencing study,^9^ we included two gene sets: SCHEMA_10 (10 genes with exome-wide significance, p < 2.5×10⁻⁶), and SCHEMA_32 (32 FDR-significant, q < 0.05), capturing the strongest rare variant associations to schizophrenia to date. We also analyzed three PPI modules derived from excitatory neuron-specific (ExN) networks in human induced neurons, prioritized based on multi-layer enrichment across rare and common variant associations, transcriptomic, proteomic, and phosphoproteomic data.^28^ Two modules were centered on HCN4 and GRIA3 (HCN4_ExN, GRIA3_ExN), while the third (SCZ_Module3) merged networks anchored on five schizophrenia-linked genes (SRRM2, SETD1A, TRIO, RB1CC1, AKAP11). As a control background, we included the WholeProteome set (∼8,000 proteins), representing baseline protein expression in neurons, which serves as a neuronal proteome background against which enrichment of genetic signal in PPI modules is typically tested.^30^ Together, these gene sets span the strongest rare variant evidence for schizophrenia as well as genetically anchored synaptic networks, allowing us to probe whether clinical subtypes show differing rare variant burdens across mechanistically relevant biological pathways.

### Computation of principal components

To account for population structure, we computed principal components (PCs) from a quality-controlled subset of the merged genetic dataset of iPSYCH, containing the iPSYCH1 and iPSYCH2 cohorts. We used imputed genotype data from 107,766 unrelated individuals of European ancestry. SNP-level quality control steps included the removal of variants with minor allele frequency (MAF) < 0.01, imputation origin p-values < 0.0001, and duplicate variant identifiers. Regions of extended linkage disequilibrium (LD) were removed, resulting in 208,928 SNPs. Additional filtering was applied thresholds of MAF ≥ 0.05, missingness rate (--geno) ≤ 0.02, and Hardy-Weinberg equilibrium (--hwe) p-value ≥ 1×10⁻⁶, leading to 185,372 SNPs. Linkage disequilibrium pruning was performed using a 500 kb window and an r² threshold of 0.5 (--indep-pairwise 500kb 0.5), reducing the set to 99,456 approximately independent SNPs. Principal component analysis was then conducted using PLINK v2.0 with the --pca approx flag to compute the first 10 principal components.

### Statistical tests

To test whether clinical subgroups were enriched for specific variables (cluster profiles, PGSs, or CADD variant counts), we used regression models matched to each outcome type. Binary outcomes (i.e. mental disorders diagnoses, attempted suicide and smoking during pregnancy) were analyzed using logistic regression; continuous outcomes (i.e birth variables and PGSs) with linear regression; and count outcomes (i.e. hospital contacts, family history and medical conditions, and number of CADD alternate alleles) with Poisson regression. If overdispersion was detected (p < 0.05 via dispersion test), we used negative binomial models (NB) instead. Zero-inflated models were not needed, as no outcome suffered from a significant excess of zeros. For each test, we created a binary indicator for cluster membership and adjusted for sex, age, iPSYCH1 membership, and the first 10 genetic PCs. Comparisons were made between each cluster and the rest combined. We excluded comparisons with <5 events for logistic and count models. Effect sizes were reported as odds ratios (ORs) for logistic models, regression coefficients (beta) for linear models, and rate ratios (RRs) for Poisson models. Multiple testing correction was applied separately for clinical, PGS, and rare variant analyses using Bonferroni and FDR. Unless otherwise noted, all plots indicate statistical significance based on Bonferroni correction within the respective analysis. For the PGS results, we additionally reported associations that were significant after FDR adjustment but not Bonferroni, to capture potentially meaningful signals that may be missed by more conservative thresholds (see results and **Fig 3A**). All tests were run in R v4.3.1^78^ using the functions `glm` (base R) or `glm.nb` (MASS v7.3-60) for NB models.^79^

## DATA AVAILABILITY

The iPSYCH data were securely stored and analyzed on the Danish national high-performance computing infrastructure (https://genome.au.dk/). All personal identifiers were anonymized prior to research use. The study was approved by the Danish Data Protection Agency, and, under Danish law, individual informed consent was not required. Access to iPSYCH data is governed by Danish regulations, and the iPSYCH consortium facilitates data access for qualified researchers in accordance with these legal frameworks. The code used in this project is available at https://github.com/leocob/SSD_RWE_paper.

## SUPPLEMENTARY DATA

Supplementary figures and tables are available in the attached files.

## AUTHOR CONTRIBUTIONS

Conceptualization: S.R, K.L, T.W. Formal Analysis: L.C. Investigation: L.C. Methodology: L.C., M.P.A, H.W., R.H.M. Resources: M.V., K.L.G.H., Y.H.H., G.P., T.W., K.L. Software: L.C., M.P.A, H.W., R.H.M. Supervision: S.R., K.L, T.W., A.R. Visualization: L.C. Writing – original draft: L.C. Writing – review & editing: L.C., S.R, Y.H.H, G.P, K.L., T.W.

