## Supplementary Figures for "Deep learning-based stratification of Schizophrenia Spectrum Disorder from real-world data reveals distinct profiles of common and rare variant genetic signal"

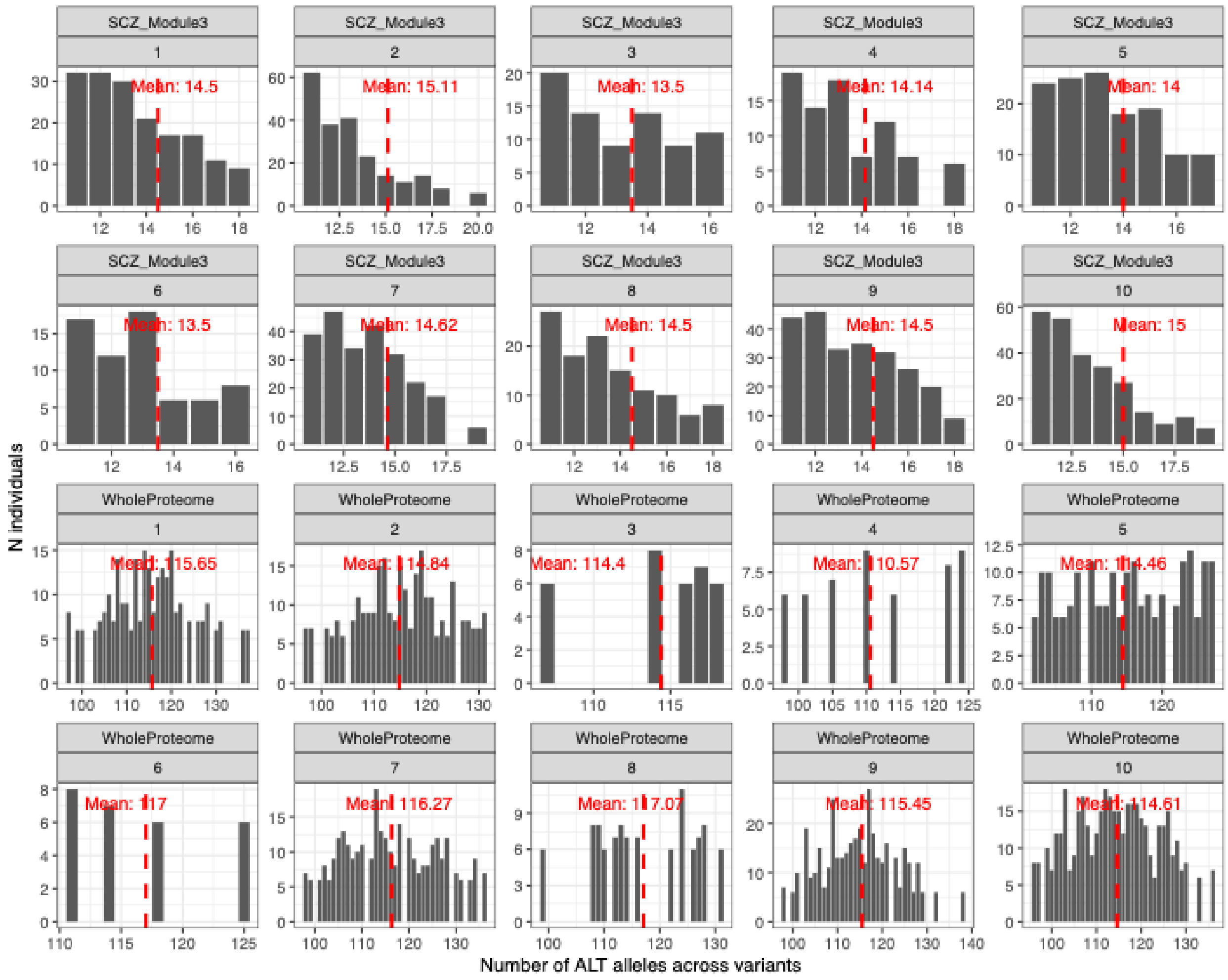


**Figure S1. Summary statistics of rare variant burden across SSD subgroups and protein networks.** Barplots and density plots showing the distribution of alternate allele counts for CADD >15 variants across the ten SSD clusters and 2 protein sets: SCZ_Module3 and WholeProteome. Note that for GDPR reasons we were not allowed to export from the iPSYCH secure cloud and show summary statistics computed on less than 5 individuals. As no individual had more than 10 alternate alleles across the gene sets SCHEMA_10, SCHEMA_32, HCN4_ExN, and GRIA3_ExN, these were not shown. Additionally, the histograms of SCZ_Module3 for number of alternate alleles < 10 are not shown.

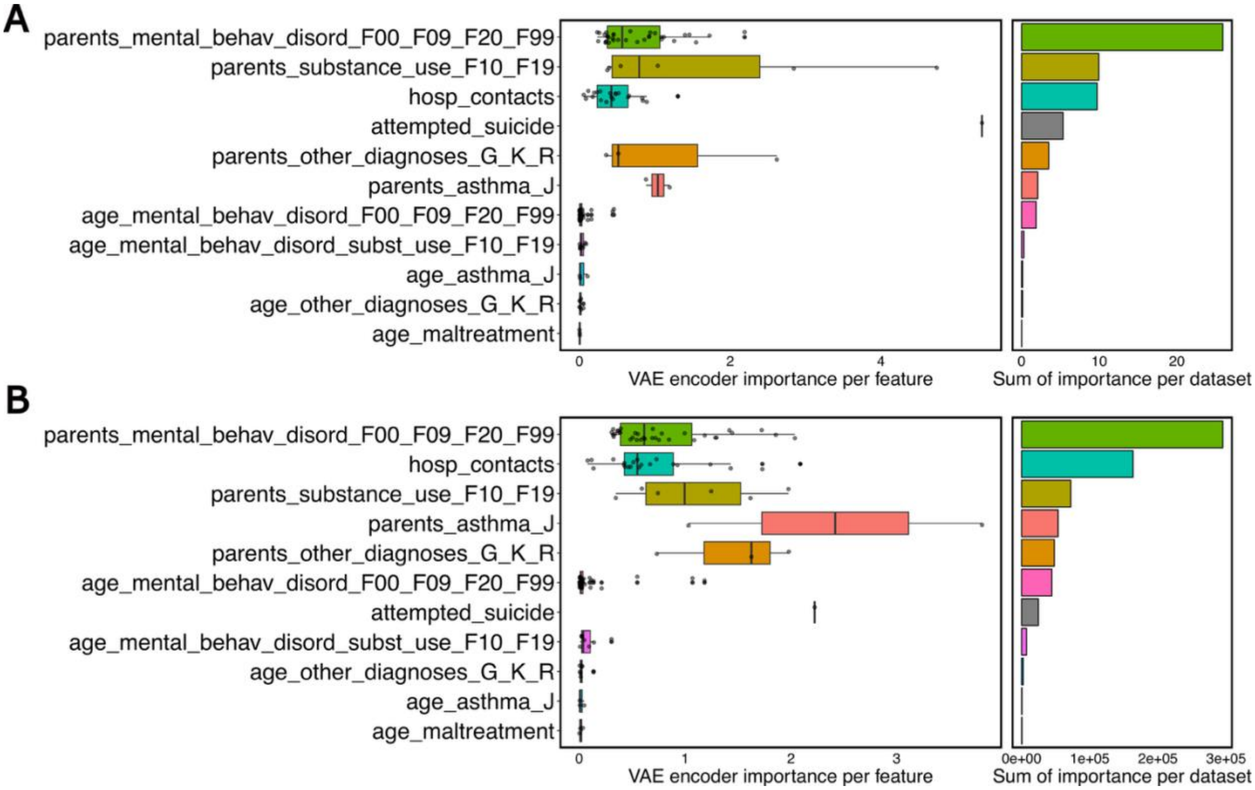


**Figure S2. Feature importance in shaping the latent space learned by the VAE. A)** Feature importance scores derived from perturbation analysis on the VAE latent space trained on the full dataset (SSD cases and controls). Top-ranking features included parental mental and substance use disorders, number of psychiatric hospital contacts, and history of suicide attempts, highlighting their influence on latent representations. **B)** Importance scores from the VAE trained on SSD cases only. While similar features remained influential, there was greater relative contribution from individual-level hospital contacts.

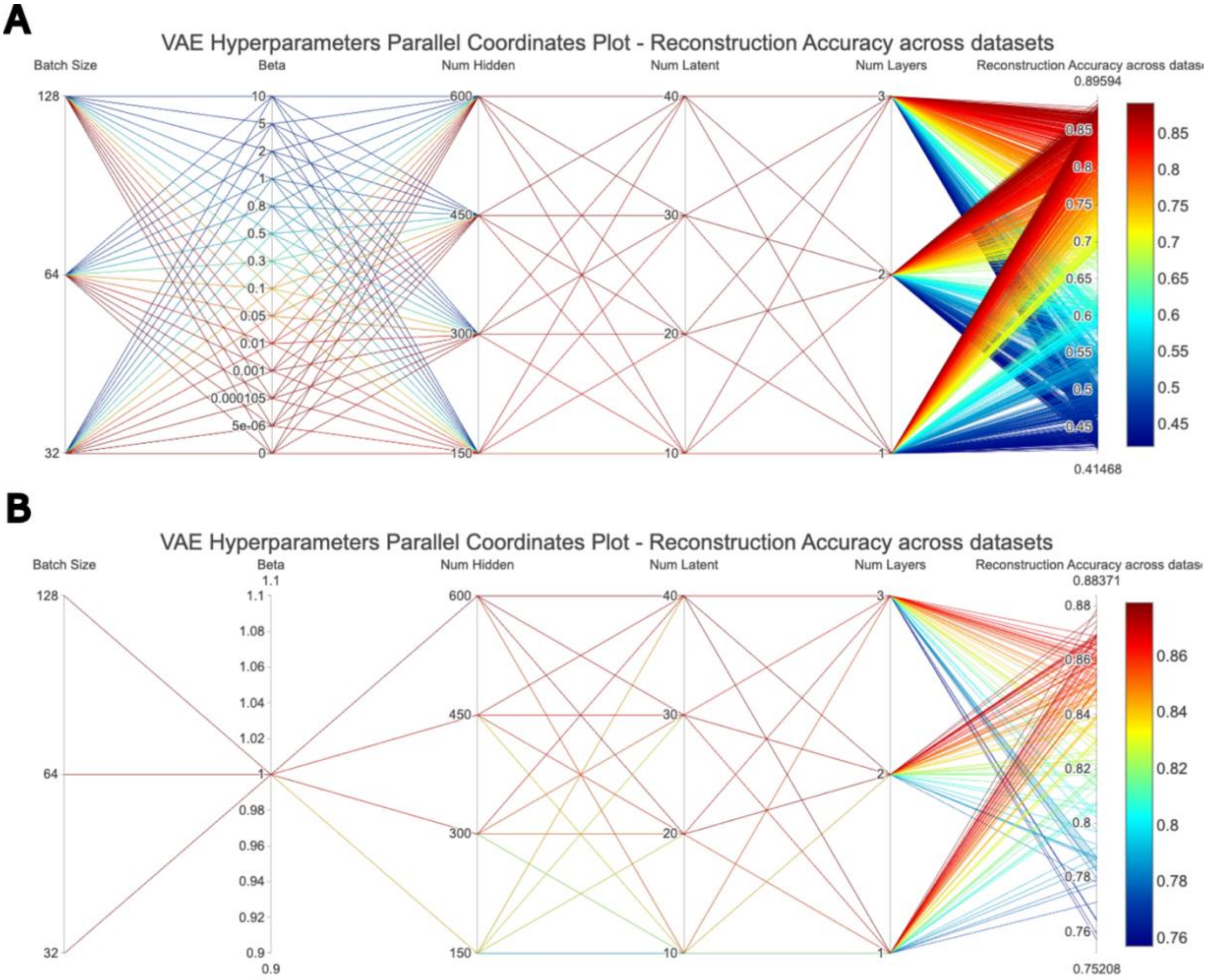


**Figure S3. Hyperparameter grid search for VAE model optimization. A)** Parallel coordinate plot of the full grid search across hyperparameter combinations tested on a random 11,000-subject subset of the full 22,092 dataset. Parameters varied included batch size (32, 64, 128), β (KL-divergence weight: 0, 5×10**−6, 1,05x10-4**, 0.001, 0.01, 0.05, 0.1, 0.3, 0.5, 0.8, 1, 2, 5, or 10), number of hidden layer units (150, 300, 450, 600), number of latent dimensions (10, 20, 30, 40), and number of hidden layers (1, 2, 3). Lines are colored by overall training reconstruction accuracy across datasets. β = 0.001 was identified as optimal, representing the highest reconstruction accuracy prior to over-regularization effects (see Fig S2). **B)** Focused subset of the grid for β value = 0.001. Reconstruction accuracy was computed as the mean cosine similarity (continuous) and classification accuracy (categorical) across the training set.

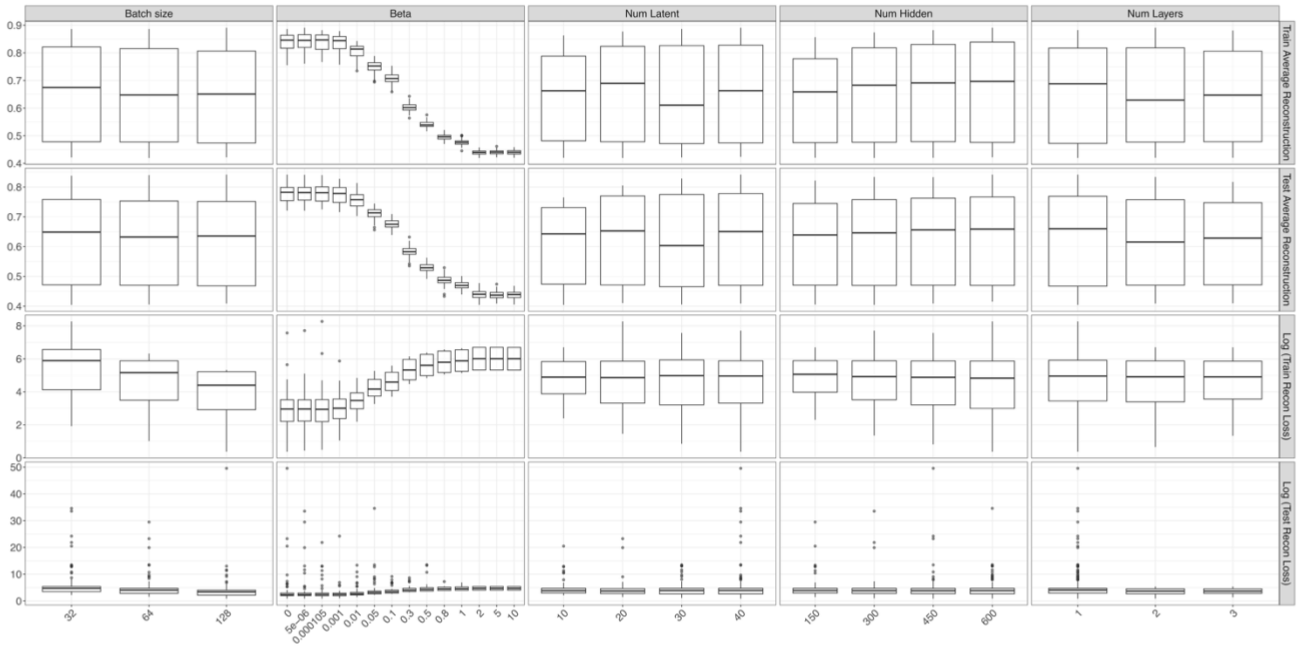


**Figure S4. Pairwise relationships between hyperparameters and model performance metrics.** Each panel shows the distribution of a performance metric (rows) across values of a specific hyperparameter at a time (columns). The hyperparameters tuned are Batch size**,** β (KL divergence weight), Number of latent dimensions**,** Number of hidden neurons, and Number of layers**.** The metrics include Train Average Reconstruction**,** Test Average Reconstruction**,** Log(Train Reconstruction Loss), and Log(Test Reconstruction Loss). Boxplots summarize performance over all runs at each hyperparameter value. Notably, the β parameter shows a strong nonlinear effect on reconstruction loss and likelihood, with optimal performance peaking around β = 0.001 before degradation is observed. We selected this value of β for all subsequent hyperparameter tuning decisions and models.

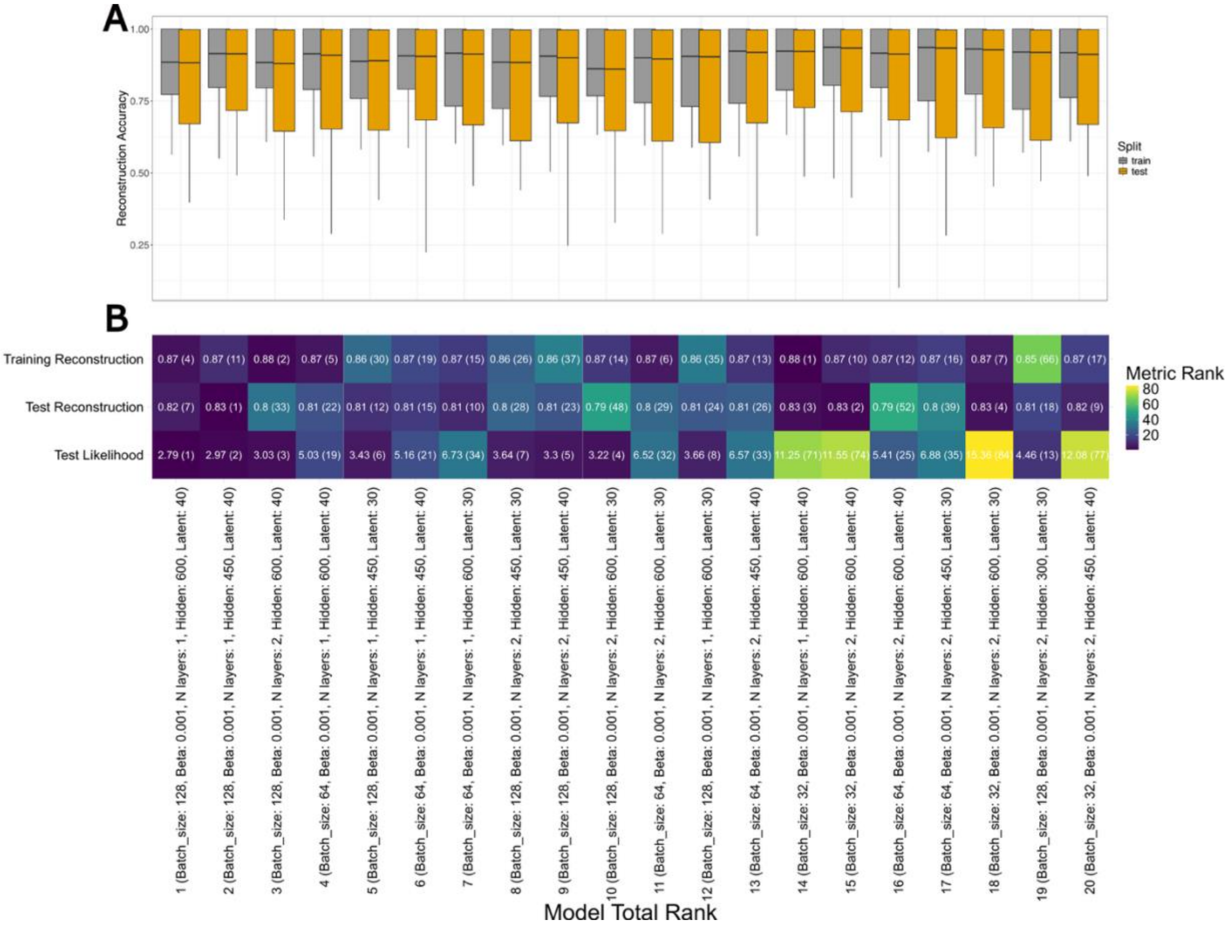


**Figure S5. Selection of top-performing VAE models based on reconstruction accuracy and likelihood. A)** Distribution of the reconstruction accuracy for the top 20 VAE models with β = 0.001 across training and test sets shown in gray and yellow, respectively. Models were trained on a random subset of the dataset (n = 11,000) with stratified 80/20 train-test splits. Models are ordered by overall rank, calculated by summing individual ranks across the three metrics, as shown in panel B. **B)** Model ranking process: models were scored by their average rank across three metrics, average reconstruction accuracy across training and test sets, and test log-likelihood. Each model's rank in individual metrics is shown in parentheses. Lower total rank indicates better overall performance.

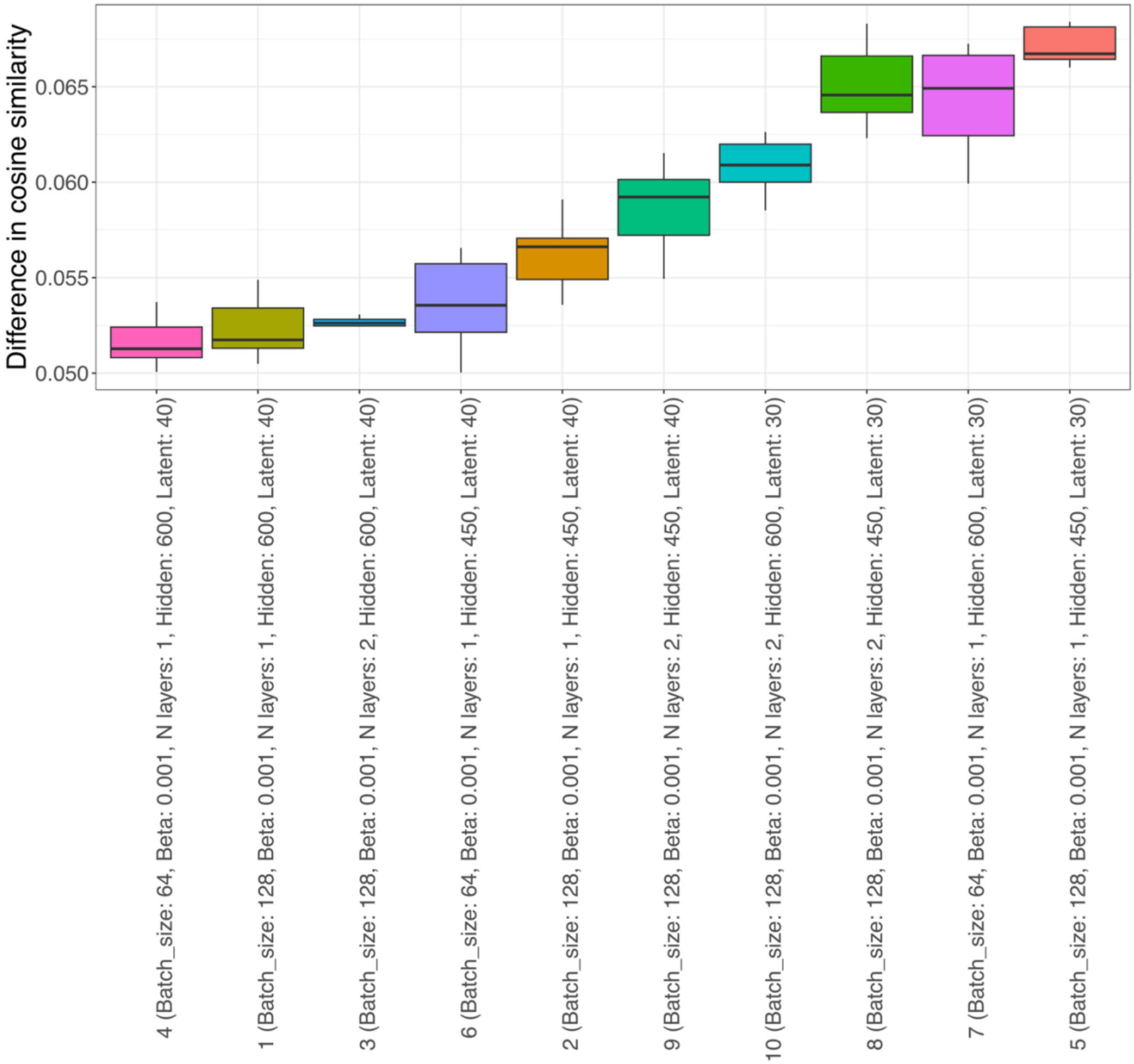


**Figure S6. Stability assessment of top VAE models using latent space similarity.** Boxplot showing the average difference in cosine similarity between latent space representations across 10 repeated training runs for each of the top 10 VAE models (selected in **Figure S3**). Lower values indicate higher stability in how individuals are embedded in the latent space across different model initializations. Model #4 (far left), defined by a batch size of 64, β = 0.001, 1 hidden layer with 600 units, and 40 latent dimensions, showed the smallest variation in cosine similarity, indicating the highest stability. This model was therefore chosen for all downstream analyses.

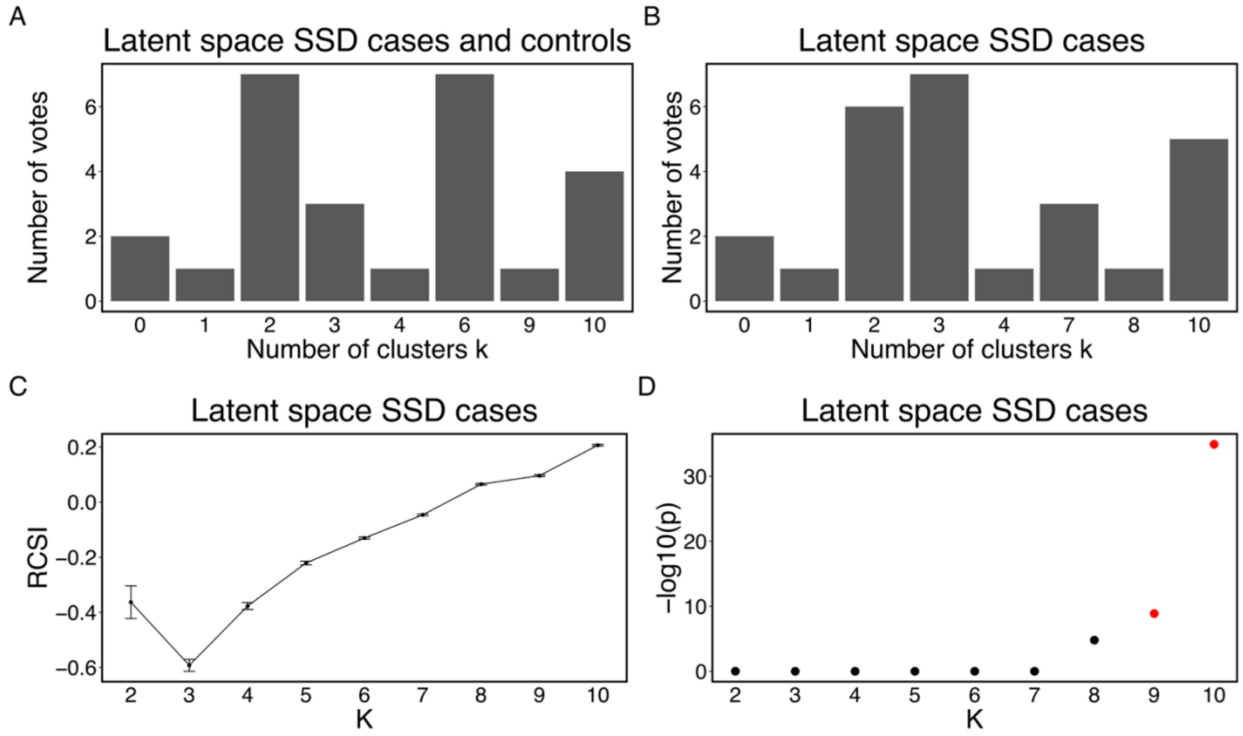


**Figure S7 Determining the optimal number of clusters for patient stratification. A)** Cluster number support based on 26 internal validation indices from NbClust for the full sample of SSD cases and controls. A two-cluster solution was most strongly supported, and this resulted in clusters A control-like and cluster B (SSD-enriched). **B)** NbClust results for SSD cases only, revealing a more complex clustering structure, with highest support for k = 3, 2, and 10. **C)** Relative cluster stability index (RCSI) from Monte Carlo reference-based consensus clustering (M3C), comparing observed versus null clustering stability. **D)** Corresponding $-\log_{10}$(p-value) for each k from M3C. A statistically significant peak was observed at k = 10, supporting this solution as the most stable and non-random stratification for within-SSD clustering.

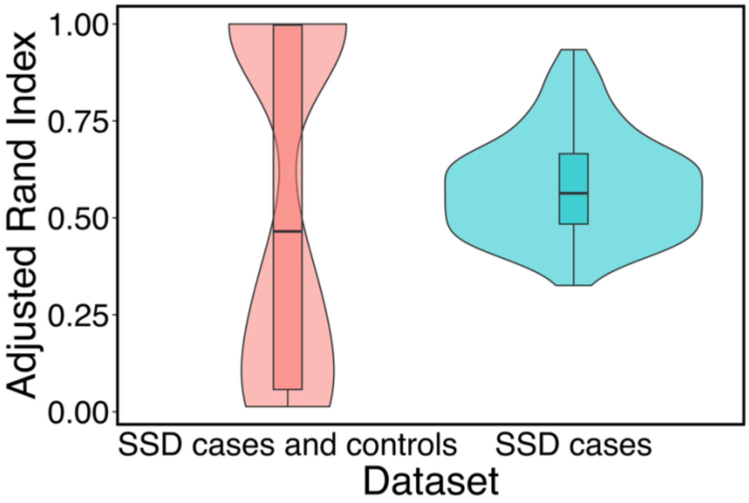


**Figure S8. Clustering stability across VAE latent representations.** Boxplots showing the distribution of Adjusted Rand Index (ARI) values across 100 repeated runs of k-means clustering. Stability was higher for clustering performed on SSD cases only (median ARI = 0.56) compared to the full dataset of SSD cases and controls (median ARI = 0.46). This suggests greater robustness in subgrouping within clinically homogeneous populations.
